# Causal relationship between school-entry age and adolescent health: quasi-experimental evidence from 1.6 million participants in Vietnam

**DOI:** 10.1101/2024.09.18.24313904

**Authors:** Ishaan Busireddy, Janny Liao, Hoa T. Nguyen, Vu Dat, Tam Tran Ngo Minh, Duc Le Thuc, Jan-Walter De Neve

## Abstract

**Background:** School-entry age has been suggested to affect human capital development. Little is known, however, about the impacts of school-entry age on adolescent health in low- and middle-income countries where most children and adolescents worldwide reside.

**Methods:** Data on children’s outcomes were extracted from the longitudinal Young Lives Study in Vietnam, conducted between 2001 and 2016 (*N*=1,532), and the Vietnam Population and Housing Censuses of 1989, 1999, and 2009 (*N*=1,595,365). In Vietnam, children need to turn six years old by December 31^st^ to enter Grade 1 in September that school year. As a result of the school-entry age policy, children born on or before December 31^st^ start school one year earlier compared to children who are born just after December 31^st^. Using exposure to the policy as an exogenous instrument for school-entry age, we used two-stage least squares regressions to determine the causal effect of school-entry age on education and health outcomes by age 23 years. We considered pre-primary education, school attendance, time spent in school, childbearing, marriage or cohabitation, as well as anthropometric measurements.

**Results:** Children born after the school-entry age cut-off were more likely to have participated in pre-primary education and were more likely to be in school when compared to children born before the cut-off. A one-year increase in age at the beginning of Grade 1 because of the policy was associated with an increase in the probability of pre-primary education of 13 percentage points (95% CI: 7.8-18.5), daily time spent in school of 0.9 hours (95% CI: 0.5-1.4) and a reduced probability of having an own child of 25.3 percentage points (95% CI: 4.4-46.2). We observed a qualitatively smaller and statistically non-significant relationship between school-entry age and measured body mass index. These results were generally consistent when using alternative specifications of our model, sample, survey rounds, and data sources.

**Discussion:** Children who are older when entering primary school stayed in school longer and postponed family formation compared to children who entered school earlier because of the school-entry age policy in Vietnam. Relative age for grade should be considered when designing sexual and reproductive health interventions and programs targeted to adolescents.

## Introduction

Most governments mandate age thresholds to enter public school. These policies set up a threshold rule for enrollment eligibility into Grade 1 of primary school. Due to the use of age cutoff dates, the oldest children are on average one year older compared to the youngest children in their Grade 1 cohort ^1^. The impacts of these differences in relative age hinges on individual development and context. On the one hand, earlier entry into school might offer cognitive benefits and accelerate social interactions and integration. Earlier school-entry may also increase exposure to school-based health programs (such as daily meals ^2,3^ or mental health screening ^4^) and offset expenditures for childcare among their parents ^5–7^.On the other hand, later entry into school may foster maturity and reduce vulnerability to negative school exposures (such as bullying) and peer pressure to engage in risky health behaviors (such as unprotected early sexual intercourse or substance use) ^8^. Late starters can also spend more time in early childhood education programs. In Vietnam, a one year increase in early childhood education has been suggested to improve literacy and numeracy skills in adolescence by 20-30 percentage points, respectively ^9^. Empirical research is therefore needed to understand the full range of impacts of school-entry cut-off dates on health outcomes and how these impacts impact life course trajectories in different contexts.

The current understanding of the impacts of school-entry age, however, largely stems from research in high-income settings such as the United States ^10^, Germany ^11^, and Denmark ^8^. This creates a crucial knowledge gap in low- and middle-income countries (LMICs), where educational systems, socio-economic factors, and cultural values differ considerably. Applying findings from high-income settings to LMICs might be misleading, as factors like access to quality pre-school programs, the nature of academic pressure, and family support can vary greatly. Therefore, research specifically exploring the context of LMICs is essential to understand the unique implications of school-entry age on adolescent health in lower-resource settings. A recent systematic review on the impacts of school-entry age on health identified only a handful of studies from LMICs ^12^. Moreover, most studies used observational designs and may be vulnerable to residual confounding or reverse causality ^13,14^. Additionally, data on age at the beginning of Grade 1 is limited, including in commonly used household- or school-based surveys which are focused on children ^15^ and adolescents ^16^. Due to these data limitations, prior studies have typically focused on using date of birth to determine the impacts of school-entry eligibility (based on month of birth) rather than using actual observed school-entry age of adolescents ^10,17^.

A randomized controlled trial, however, would be costly, take many years to conduct, and require an ethically questionable random assignment of individuals to different study arms where some children would need to enter school early (late). In the absence of such a randomized experiment, natural experiments offer a valuable opportunity for causal inference in population health ^18,19^. In this study, we therefore employ a quasi-experimental approach which leverages a long-standing education policy in Vietnam. The basic framework underpinning causality in our study is like that of a randomized controlled trial where participants are assigned to a treatment and control group. Specifically, we exploit the natural variation in school-entry ages due to Vietnam’s enrollment cut-off dates ^20^. In Vietnam, a school-entry age cut-off based on month of birth delays the school-entry age of children who were born after the cut-off (treatment group) compared to children who were born before the cut-off (control group). Because parents have limited control over the exact timing of birth of their children, we can compare the outcomes of adolescents as if they were randomized into treatment and control groups in childhood by the education policy ^21,22^. We determined the impacts of school-entry age on several adolescent outcomes, including pre-primary education, school attendance, childbearing, marriage, as well as anthropometric measurements, using data from the Young Lives Study, which followed thousands of children from early childhood up until age 23 years, and the Vietnam Population and Housing Censuses of 1989, 1999, and 2009. This study aims to bridge the knowledge gap and provide robust evidence for Vietnamese policymakers, ultimately guiding interventions towards reducing developmental inequities that foster both academic success and positive health trajectories for Vietnamese youth.

## Methods

### Data sources and study population

#### Young Lives Study

Longitudinal data on socio-demographic and health outcomes were extracted from the Young Lives data of Vietnam. This dataset contains information on about 3,000 children who have been surveyed once every 3-4 years since 2001 ^23^. Round 1 of the study surveyed two birth cohorts of children, including 1-year-olds (born in 2001-02) and 5-year-olds (born 1994-95). Round 5 surveyed them when they were between 15 and 23 years old. The younger children were tracked from infancy to their mid-teens and the older children were tracked through adulthood, when some became parents themselves. Data was collected from families and directly from the children themselves. Strengths of the Young Lives data include the availability of data on early life exposures, such as children’s age at the start of the school year in Vietnam, and the ability to link these early life exposures to measured educational and health outcomes up until early adulthood, such as measured body mass index. To our knowledge, no other datasets from LMICs include data on school-entry age and measured adolescent health outcomes. Attrition rates in the Young Lives data are low. Over 90% of children in Round 1 were followed-up in Round 5 ^23^. Information on exact date of birth was classified as protected personal data. We therefore estimated children’s month of birth based on their date of interview (variable *dint*) and children’s age in months (variable *agemon*). We limited the study population to children with complete data on school-entry age and anthropometric measurements in the latest round of the Young Lives data (Round 5). We limited the sample to children who were born 3 months before and after the school-entry age cutoff (Dec 31^st^) to maximize the comparability of early starters and late starters on observed and unobserved characteristics, yielding a final sample of 1,334 respondents born between October and March. In supplementary analyses, we used alternative rounds of the Young Lives study to assess whether the observed relationship was consistent across different periods. We also included smaller and larger windows of months of birth around the cut-off (2 and 5 months).

#### Vietnam Population and Housing Census

To explore whether our findings may generalize to Vietnam more broadly, we also extracted data from the Vietnam Population and Housing Censuses of 1989, 1999, and 2009 through the Integrated Public Use Microdata Series (IPUMS) ^24^. The Censuses were conducted by the Bureau of the Central Steering Committee, General Statistics Office, Vietnam, using a systematic stratified sampling to create random 5% (Census 1989) and 3% (Census 1999), and 15% (Census 2009) samples of the population universe. Similar to our study population from the Young Lives Study, we limited the sample to all respondents who were born 3 months before and after the school-entry age cutoff (Dec 31^st^) to maximize the comparability of early starters and late starters. Data on month of birth, demographics, school attendance, childbearing, and marital status were available for 99% of eligible respondents ages 15-23 years, yielding a total sample of 1,595,365 individuals born between October and March. IPUMS harmonizes variables across Censuses so that the same codes have the same meaning across all Censuses. Additional details on these data sources and study population are available in **Text S1** in the Appendix.

### Exposure

Our key exposure was age at primary school entry (in years). School-entry age was defined as “Child’s age at start of Grade 1” and was calculated by the Young Lives study team using the participants’ exact date of birth and the start of the academic year in Vietnam (September) for children who were enrolled in Grade 1. All adolescents in our analytical sample had attended at least some primary school by age 15 - 23 years and thus had data on our exposure.

### Exogenous instrument

In Vietnam, Education Law stipulates that children start school in September of the calendar year in which they turn six years of age ^20^. Children therefore need to turn six years old by the cut-off of December 31^st^ to enter Grade 1 in September that school year. The school year starts in the first week of September and runs until the end of May the following calendar year. Primary schooling in Vietnam is compulsory and universal primary education has been achieved ^25^. As a result of the school-entry age policy, children who are born just before December 31^st^ start school one year earlier compared to children who are born just after December 31^st^. The policy has been suggested to be in place since at least 1945 when the nation became independent. To use the school-entry age policy as a natural experiment, we defined a variable that takes the value one if a child is born after the school entry cut-off date (January – March); and zero if the child is born before the cut-off date (October – December). Following prior literature on the impacts of school-entry age, we normalize month of birth as the number of months before and after the cut-off ^17,26^. Additional information on the education system and context is presented in **Text S2**.

### Control variables

Accounting for control variables in our analysis of the education policy may reduce the variance and mitigate small biases when including observations that are further apart from the December 31^st^ threshold. In our analyses using Young Lives data, we controlled for age (continuously in years) and an indicator for gender. In supplementary analyses, we also ran models without control variables as well as when controlling for additional control variables such as indicators for age (as opposed to age continuously in years), a continuous linear term in year of birth, a continuous linear term in month of birth, as well as an indicator for Young Lives birth cohort, including the younger (born 2001-02) and older Young Lives birth cohorts (born 1994-95). When using the Census data, we controlled for age (years) and indicators for census year (period effects). The availability of several Censuses allowed us to generate variation in age for a given birth cohort. By simultaneously controlling for age and Census year, we therefore also controlled for birth cohort effects (since birth cohort equals Census year *minus* a respondent’s age).

### Outcome measures

Our main outcomes were ever attended pre-primary education or not (binary), current school attendance (binary), school participation (number of hours in school recorded based on a typical day), ever having had a child (binary), ever having married or cohabited (binary), and measured body mass index (BMI) (kg/m^2^). Height and weight were measured twice standing up on a weighing scale and height board wearing only light clothes. If there was a large difference between two measurements, children were measured one more time and the most common measurement was recorded. Those health outcomes were chosen based on prior literature on the relationship between relative age for grade and sexual and reproductive health ^17,27^ and nutritional outcomes ^28^ in the context of LMICs. Data on education and measured BMI were available for both the younger (aged 15-16 years) and older birth cohorts (aged 21-23 years) of the Young Lives study. Data on childbearing and marriage or cohabitation was only available for the older birth cohorts in the Young Lives Study. Our outcomes in the Census data were school attendance, number of children ever born (women), number of own children living in the household (men), and marriage or cohabitation. Data on other relevant health outcomes were not available in the Census data.

### Statistical analyses

We analyzed the school eligibility cut-off in three steps. First, we determined whether cohorts who were born just after the December 31^st^ cut-off had a higher school-entry age compared to cohorts who were born just before the cut-off (“first stage”). We estimated the impact of the eligibility cut-off on school-entry age using multivariable ordinary least squares (OLS) regression models. To assess the robustness of our first stage, we used several model specifications, including without covariates; a continuous term in age and gender; age, gender, and month of birth (continuously); indicators for single-year age groups, gender, and month of birth (continuously); and year of birth, gender, and month of birth (continuously). Second, we assessed the intention-to-treat (ITT) effect of being born after the school-entry age eligibility cut-off on our education and health outcomes in multivariable linear probability models. Natural experiments altering exposure probabilities can be analyzed akin to randomized controlled trials with non-compliance ^21^. Third, under additional assumptions, we can scale our ITT estimates by the first stage estimates above to obtain the local average treatment effect of age-at-entry ^29^. To do so, we employed two-stage least squares (2SLS) regression models using exposure to the school-entry age cutoff as an instrumental variable (IV) for school-entry age while adjusting for covariates. We assume that our IV estimates are local to the subpopulation who complied with their treatment assignment, meaning that these children experience increased school-entry age as a result of the school eligibility cut-off ^29^.

### Supplementary analyses

In addition to the sensitivity analyses described above, we conducted several additional analyses to generate further confidence in the robustness of our results. First, one possible threat to the validity of our results may be seasonality in births or manipulation in month birth around the cutoff. Causal inferences on an exposure based on month of birth could be undermined by selection based on parental characteristics, manipulation of birth dates, or early-life exposures. Sophisticated parents, for example, may time the birth of their children based on the school-entry age cutoff ^30^. To rule out manipulation of birth dates, we therefore show the distribution in months of birth around the cutoff. To rule out selection and confounding by early life exposures, we also assess for balance at the threshold in parental characteristics. We plot maternal characteristics by children’s month of birth, including maternal educational attainment, maternal age, antenatal care (number of antenatal care visits and vaccinations) as well as children’s birthweight (grams). Second, to further explore the robustness of our results, we assess the relationship between school-entry age and month of birth (first stage) separately by gender and area of residence. Third, we replicated our ITT results using data from several rounds of the Vietnam Census, controlling for covariates, and determined heterogeneity by children’s age and gender by running our regression models separately for each gender and single-year age groups.

### Code and ethical clearance

All analyses were conducted using Stata MP v.17. The study was pre-registered and approved by the Heidelberg University Hospital Ethics Committee (S-825/2022).

## Results

### Descriptive statistics

**Table 1** lists all data sources and sample specifications. In **Tables S1 – S2**, we show selected characteristics of our analytical samples. Our final sample from the Young Lives Study included 1,334 respondents aged between 15 and 23 years old who were born between October and March. Out of those respondents, 908 children (68.1%) were part of the Young Lives younger cohort born between 2001 and 2002, and 426 children (31.9%) were part of the Young Lives older cohort born between 1994 and 1995. The average age in our sample was 17.3 years old and average age at beginning of Grade 1 was 5.4 years old implying that the average duration between our exposure (school-entry age) and our educational and health outcomes was about 12 years. Although the majority of children resided in rural areas (85.2%), most attended at least some pre-primary education (70.0%). Among children in the younger cohort, 763 children (84.0%) were still enrolled in school at the time of the Round 5 survey whereas among children in the older cohort only 88 children (20.1%) were still enrolled. Among children in the older cohort, 122 children (28.6%) had been married or cohabited and 90 children (21.1%) had a child themselves (both genders). The average BMI in the pooled sample was 19.5 kg/m^2^ and ranged from 12.6 to 40.5 kg/m^2^. In the younger cohort, 103 children (11.3%) were stunted (defined as having height-for-age z-scores less than two standard deviations below the median of a reference population of the same age and gender) and 167 children (9.0%) suffered from overweight or obesity (defined as having a BMI-for-age z-score of one above the median of the reference group). In the Census data, average age was 18.7 years old and 69.9% of respondents resided in rural areas (**Table S2**).

**Table 1.** Data sources and sample specifications.

| *Data source* | **Year** | **Sample** | **N** |
| --- | --- | --- | --- |
| Young Lives cohorts, Round 5 | 2016-17 | Ages 14-23 | 1,382 |
| Young Lives cohorts, Round 4 | 2013-14 | Ages 11-20 | 1,364 |
| Young Lives cohorts, Round 3 | 2009-10 | Ages 7-16 | 1,460 |
| Young Lives cohorts, Round 2 | 2006-07 | Ages 4-13 | 1,476 |
| Young Lives cohorts, Round 1 | 2002 | Ages 0-9 | 1,532 |
| Census, 15% sample | 2009 | Ages 15-23 | 1,185,080 |
| Census, 3% sample | 1999 | Ages 15-23 | 201,698 |
| Census, 5% sample | 1989 | Ages 15-23 | 208,587 |

*Notes:* Data sources used in the current study. Sample size includes all respondents who were born between October and March (3 months before and after the school-entry age cutoff).

### Exogenous instrument: school-entry age by month of birth

**Figure 1** shows entry into primary school by month of birth in Vietnam. In Figure 1(a), we show the probability of attending Grade 1 by month of birth among children aged 6 years using Census data. Consistent with the Education Law in Vietnam, there is a discontinuity in the probability of enrollment in Grade 1 by month of birth. Children who were born just after the December 31^st^ cutoff were on average less likely to be in school compared to children who were born just before the cut-off. In Figure 1(b), we show actual (observed) school-entry age by month of birth using data from the Young Lives Study for the same birth cohorts (born after 1992). Children born between January and March are on average 5.7 years old whereas children born between October and December are on average 5.0 years old, a difference of about 0.7 years. Moreover, we observe similar jumps in entry into primary school for the different Young Lives cohorts and when using different Census rounds, suggesting that the policy was indeed a consistent feature of the education system in Vietnam. We note that the difference in school-entry age shown in Figure 1b is less than one year because some children born January-March start primary school a year early when they are five and some children enter primary school late despite the legal requirement. The relationship between being born after December 31^st^ and school-entry age persisted when controlling for a wide range of control variables, including when adding indicators for age, year of birth continuously, month of birth continuously, as well as an indicator for Young Lives birth cohort (**Table 2**). In multivariable OLS models, being born between January and March was associated with an increase in school-entry age between 0.5 and 0.8 years in the Young Lives data.

**Figure 1.**
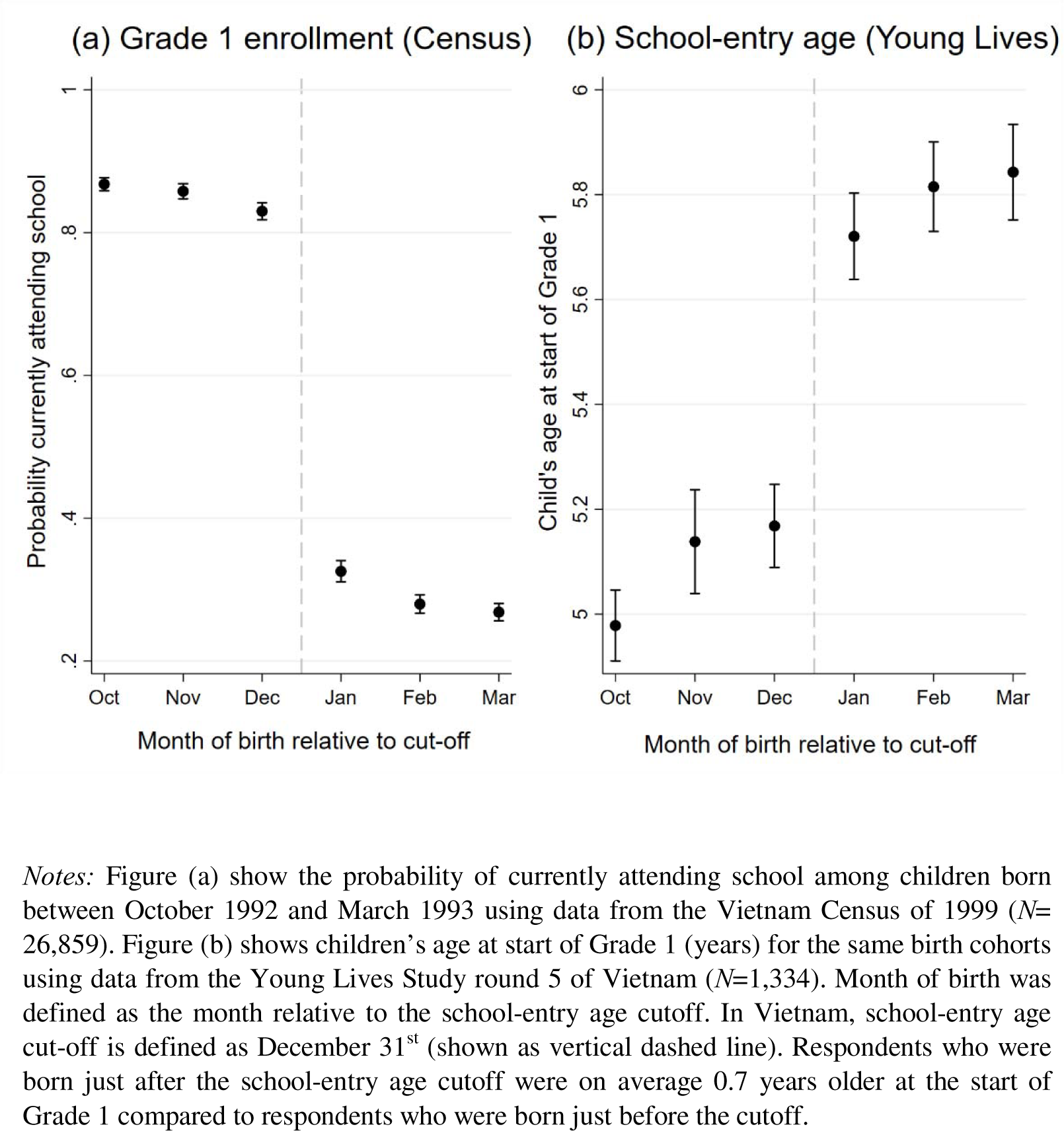
Entry into primary school in Vietnam.

**Table 2.** First-stage OLS regression results: the relationship between being born after December 31^st^ and school-entry age among adolescents and youth aged 15 - 23 years old (Young Lives)

|  | (1) | (2) | (3) | (4) | (5) |
| --- | --- | --- | --- | --- | --- |
| *Dependent variable (DV):*<br>*Age at start of grade 1 (years)* | Without<br>control<br>variables | Adding age<br>and gender | Adding age,<br>gender,<br>month of<br>birth | Adding age<br>indicators,<br>gender,<br>month of<br>birth | Adding year<br>of birth,<br>gender,<br>month of<br>birth |
| *Predictor* |  |  |  |  |  |
| Born January-March (1=yes, 0=no) | 0.698***<br>(0.035) | 0.682***<br>(0.034) | 0.473***<br>(0.069) | 0.464***<br>(0.068) | 0.794***<br>(0.085) |
| *Additional covariates* |  |  |  |  |  |
| Age (continuous) | - | ✓ | ✓ | - | - |
| Indicators for single-year age group | - | - | - | ✓ | - |
| Indicator for gender | - | ✓ | ✓ | ✓ | ✓ |
| Month of birth (continuous) | - | - | ✓ | ✓ | ✓ |
| Year of birth (continuous) | - | - | - | - | ✓ |
| Indicator for Young Lives cohort | - | - | - | - | ✓ |
| *Data source and sample* |  |  |  |  |  |
| Young Lives Study (Round 5) | ✓ | ✓ | ✓ | ✓ | ✓ |
| Born between October-March | ✓ | ✓ | ✓ | ✓ | ✓ |
| Mean DV, October-December birth cohorts | 5.1 | 5.1 | 5.1 | 5.1 | 5.1 |
| Observations | 1,334 | 1,334 | 1,334 | 1,334 | 1,334 |
| R-squared | 0.224 | 0.274 | 0.280 | 0.292 | 0.291 |

*Notes:* Table shows ordinary least regression (OLS) results for the relationship between being born after December 31<sup>st</sup> (binary) and age at school entry (years). Model 1 includes no covariates, Model 2 includes a continuous term in age and gender; Model 3 includes age, gender, and month of birth (continuous); Model 4 includes single-year indicators for age, gender, and month of birth; and Model 5 includes year of birth, month of birth, and gender. Robust standard errors are in parentheses. The sample includes all adolescents and youth ages 15-23 years in the Young Lives Study survey Round 5 conducted in 2016 ( $N=1,334$ ). \*\*\* $p<0.01$ , \*\* $p<0.05$ , \* $p<0.1$ .

### Causal effect of school-entry age on adolescent health

#### Intention-to-treat regression results

In **Table 3**, we show ITT regression results using data from the Young Lives Study and controlling for age and gender (Panel A). Being born after the cutoff was associated with a 9-percentage point increase (95% CI: 5.2-12.7) in the probability of having attended pre-primary education. Late starters were also 11.3 percentage points more likely (95% CI: 7.3-15.3) to be currently in school; spent 0.6 hours per day more (95% CI: 0.35-0.91) in school; and were 10.2 percentage points less likely (95% CI: 2.4-17.9) to have had a child by age 23. Being born after the school-entry cutoff was also associated with a qualitatively large reduction in the probability of marriage or cohabitation although this result did not reach conventional benchmarks of statistical significance in the Young Lives data (-8.2 percentage points, 95% CI: -17.0-0.4). In terms of anthropometric measurements, differences by month of birth were qualitatively small and non-significant. Our results were generally consistent across different datasets and measures. In **Table 4**, we show ITT results using data from the 1989, 1999, and 2009 Census. Being born after December 31^st^ was associated with a 4.7 percentage point increase (95% CI: 4.5-4.8) in school attendance among women and a 5.3 percentage point increase (95% CI: 5.1-5.5) among men aged 15-23 years. Women also saw a reduction of 0.052 children ever born (95% CI: 0.050-0.054). **Figure S1** in the Appendix graphically displays the unadjusted relationship between early family formation and month of birth during the period of late adolescence. Adolescents who were eligible to enter school late in childhood (born between January and March) were considerably more likely to still be attending school, had fewer children, and were less likely to have ever married or cohabited.

**Table 3.** ITT and 2SLS regression results: the relationship of school-entry age with educational and health outcomes among participants aged 15-23 years in Vietnam (Young Lives)

| *Dependent variable (DV):* | (1)<br>Child has<br>attended pre-primary<br>school<br>(1=yes) | (2)<br>Currently<br>attending<br>school<br>(1=yes) | (3)<br>Hours per<br>spent day<br>in school | (4)<br>Has son or<br>daughter<br>(1=yes) | (5)<br>Ever<br>married or<br>cohabited<br>(1=yes) | (6)<br>Body mass<br>index<br>(kg/m2) |
| --- | --- | --- | --- | --- | --- | --- |
| *Panel A. Intention-to-treat results (ITT)* |  |  |  |  |  |  |
| *Predictor* |  |  |  |  |  |  |
| Born January-March (1=yes, 0=no) | 0.090***<br>(0.019) | 0.113***<br>(0.020) | 0.632***<br>(0.144) | -0.102***<br>(0.039) | -0.083*<br>(0.044) | 0.075<br>(0.163) |
| *Additional covariates* |  |  |  |  |  |  |
| Age (years) | ✓ | ✓ | ✓ | ✓ | ✓ | ✓ |
| Gender | ✓ | ✓ | ✓ | ✓ | ✓ | ✓ |
| *Data source and sample* |  |  |  |  |  |  |
| Young Lives Study (Round 5) | ✓ | ✓ | ✓ | - | - | ✓ |
| Born between October-March | ✓ | ✓ | ✓ | ✓ | ✓ | ✓ |
| Mean DV, October-December birth cohorts | 0.669 | 0.599 | 3.7 | 0.239 | 0.303 | 19.5 |
| Observations | 1,334 | 1,334 | 1,334 | 426 | 426 | 1,334 |
| R-squared | 0.452 | 0.392 | 0.359 | 0.072 | 0.086 | 0.062 |
| *Panel B. Two-stage least squares results (2SLS)* |  |  |  |  |  |  |
| *Predictor* |  |  |  |  |  |  |
| Age at start of grade 1 (years) | 0.131***<br>(0.027) | 0.166***<br>(0.031) | 0.926***<br>(0.217) | -0.253**<br>(0.107) | -0.205*<br>(0.114) | 0.111<br>(0.239) |
| *Additional covariates* |  |  |  |  |  |  |
| Age (years) | ✓ | ✓ | ✓ | ✓ | ✓ | ✓ |
| Gender | ✓ | ✓ | ✓ | ✓ | ✓ | ✓ |
| *Data source and sample* |  |  |  |  |  |  |
| Young Lives Study (Round 5) | ✓ | ✓ | ✓ | - | - | ✓ |
| Born between October-March | ✓ | ✓ | ✓ | ✓ | ✓ | ✓ |
| F-statistic | 392.6 | 392.6 | 392.6 | 39.0 | 39.0 | 392.6 |
| Observations | 1,334 | 1,334 | 1,334 | 426 | 426 | 1,334 |

*Notes:* Panel A was an Intention-to-treat (ITT) ordinary least squares (OLS) linear probability model. Panel B was a 2-stage least squares (2SLS) linear probability model in which exposure to increased school-entry age from the school-entry age cut-off was used as an instrumental variable (IV) for the respondent's age at entry into Grade 1 (in years). All models controlled for age (continuously in single years) and gender. Sample includes respondents ages 15-23 years at the time of the survey in the Young Lives study Round 5 of Vietnam with complete data on educational outcomes and body-mass index born between October and March ( $N=1,334$ ). Data on fertility and marriage was not available for the younger birth cohorts in the Young Lives study. \*\*\* $p<0.01$ , \*\* $p<0.05$ , \* $p<0.1$

**Table 4.** ITT regression results: relationship of being born after December 31^st^ with school attendance and early family formation among participants aged 15-23 years (Census data)

| *Intention-to-treat results (ITT)* | (1) | (2) | (3) | (4) | (5) | (6) |
| --- | --- | --- | --- | --- | --- | --- |
|  | **Female** |  |  | **Male** |  |  |
| *Dependent variable (DV):* | **Currently attending school (1=yes)** | **Number of children ever born** | **Ever married or cohabited (1=yes)** | **Currently attending school (1=yes)** | **Number of own children in household** | **Ever married or cohabited (1=yes)** |
| *Predictor* |  |  |  |  |  |  |
| Born January - March (1=yes, 0=no) | 0.047***<br>(0.001) | -0.052***<br>(0.001) | -0.044***<br>(0.001) | 0.053***<br>(0.001) | -0.021***<br>(0.001) | -0.024***<br>(0.001) |
| *Additional covariates* |  |  |  |  |  |  |
| Age (years) | ✓ | ✓ | ✓ | ✓ | ✓ | ✓ |
| Census year | ✓ | ✓ | ✓ | ✓ | ✓ | ✓ |
| *Data source and sample* |  |  |  |  |  |  |
| Census 1989, 1999, 2009 | ✓ | ✓ | ✓ | ✓ | ✓ | ✓ |
| Born between October-March | ✓ | ✓ | ✓ | ✓ | ✓ | ✓ |
| Mean DV, Oct - Dec birth cohorts | 0.339 | 0.229 | 0.258 | 0.339 | 0.086 | 0.119 |
| Observations | 786,983 | 786,983 | 786,983 | 808,382 | 808,382 | 808,382 |
| R-squared | 0.255 | 0.171 | 0.228 | 0.228 | 0.071 | 0.126 |

*Notes:* Table shows results from intention-to-treat (ITT) ordinary least squares (OLS) linear probability models, separately by gender. All models controlled for age (continuously in single years) and census year. Sample includes respondents ages 15-23 years at the time of the survey in Census 1989, 1999, and 2009 with complete data on educational and childbearing outcomes and born between October and March ( $N= 1,612,204$ ). \*\*\* $p<0.01$

#### Two-stage least squares regression results

In **Table 4**, we also show our main regression results when using exposure to the school-entry age policy as an instrumental variable for age at the beginning of Grade 1 (shown in Panel B). These models show results using the pooled sample (including both the younger and older Young Lives birth cohorts) while controlling for generic trends in age and gender. School-entry age had considerable impact on human capital development. Specifically, an increase of one year in age at the beginning of Grade 1 because of the school-entry age policy increased the probability of pre-primary education by 13 percentage points (95% CI: 7.8-18.5) relative to a baseline probability of 67% among cohorts born in October to December. An increase in school-entry age also increased the probability of still being in school by 16.6 percentage points (95% CI: 10.5-22.7). Children who entered Grade 1 one year older also spent 0.9 hours more in school (95% CI: 0.5-1.4), implying that late starters spent about 25% more time in school per day compared to early starters by the time they were ages 15 – 23 years old. The effect sizes on childbearing in the older cohorts were qualitatively large. Entering Grade 1 late reduced the probability of having a child by 25.3 percentage points (95% CI: 4.4-46.2). The relationship between school-entry age and measured BMI, however, was qualitatively smaller and non-significant (0.11 kg/m^2^, 95% CI: -0.35-0.58).

### Results from supplementary analyses

**Figure S2** shows the distribution of month of birth around the cutoff using data from the Young Lives Study, separately for each Young Lives survey round and cohort. We observe no evidence of bunching at the December 31^st^ threshold. We also find no differences in maternal educational attainment (years), caregiver literacy, number of antenatal visits by mothers, number of antenatal tetanus vaccinations among mothers, or maternal age by children’s month of birth (**Figure S3**). We additionally find no differences in birthweight for children born on either side of the threshold. These findings support the validity of our identification strategy. Children born in October-December vs. January-March had similar parents, grew up in similar households, and had similar early childhood anthropometric measurements. They look similar on all assessed measures until they start primary school. In our main analysis, we limited the sample to children born 3 months before and after the cut-off using cross-sectional data from Round 5 of the Young Lives study. **Tables S3-S4** show ITT results when using alternative sample specifications, including when using either smaller and larger windows of months of birth around the cut-off, and when using other rounds of the Young Lives study. Our results are consistent across all of those alternative specifications. in **Figure S4**, we show school-entry age by month of birth when stratifying the sample by gender and area of residence (urban vs. rural). Children born in January-March had higher school-entry age across all sub-populations compared to children born in October-December. The school-entry age policy affected both genders as well as children living in different within-country geographical and socio-economic contexts, in line with the school-entry age policy, which aimed to cover the entire country. Lastly, in **Figure 2**, we show differences in early family formation respondents born October – December vs. those born January – March using data from the Census 1989, 1999, and 2009, estimated separately for each single-year age group, while controlling for indicators for survey year. The increase in childbearing for women born between January – March vs. October – December appears by late adolescence and peaks by around age 22 years. Late starters are less likely to have children and marry during adolescence, putting them at lower risk of adverse maternal and child health outcomes ^31–34^, and postpone childbearing to at least ages 23 compared to women born between October and December.

**Figure 2.**
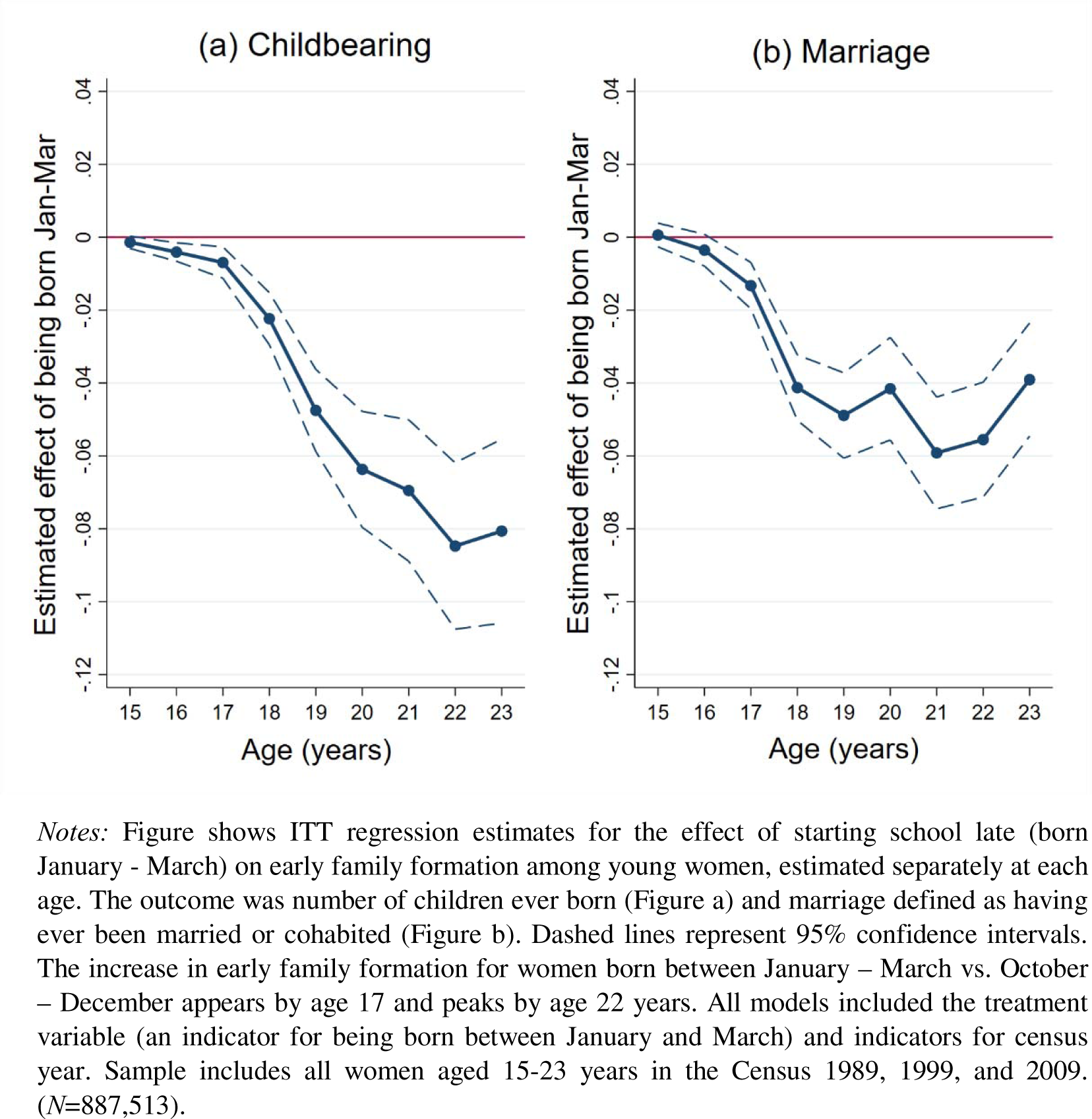
ITT regression results: early family formation by month of birth and age.

## Discussion

The age at which children enter primary school has been suggested to affect human capital development later in life, but little is known about the impacts of school-entry age on health in lower-resource settings, where most children and adolescents reside ^12^. Moreover, existing studies have employed study designs which are vulnerable to residual bias and reverse causality ^13,14,35^ and lack data on actual observed school entry age ^12^. Prior studies have relied on data on children’s date of birth from cross-sectional surveys to ascertain “eligibility” to enter school based on date of birth in relationship to a school-entry age cutoff date ^12^. Here, we sought to determine the causal relationship between school-entry age on adolescent health by triangulating several datasets from Vietnam, including data on the actual observed school-entry age of children. In doing so, a key advantage was that we were able to link an exposure in early childhood (i.e., age at the beginning of Grade 1) to health outcomes up until early adulthood ^23^. Using this dataset, we then exploited the school-entry age policy in Vietnam as a natural experiment to determine for the first time, to our knowledge, the causal relationship between actual school-entry age and several adolescent health domains in the context of LMICs. We compared the health of adolescents who were induced to start school later as a result of the school-entry age policy (akin to the “treatment group” in a randomized controlled trial) to the health of adolescents who were induced to start school earlier (akin to the “control group”) ^18^. Our results were consistent across a wide range of supplementary analyses (**Table 2** and **Tables S3-S4**). Moreover, data from the Young Lives Study included measured birthweight, and other background characteristics, which allowed us to further strengthen confidence in the key assumptions underpinning causal inference (**Figure S3**).

In any grade, there will always be some students who are the youngest; however, interventions and programs may mitigate potential detrimental impacts of school-entry age cutoffs among relatively younger students and improve the health trajectories of adolescents. Indeed, the magnitude of the effect sizes identified in our study were rather large. In the Census data, for example, ITT estimates suggested that being born after the school-entry age cutoff (January-March) was associated with a 4.4 percentage point reduction in the probability of being married among young women aged 15-23 years (**Table 4**). Given a baseline prevalence of about 25% among those born October-November, this implies a 18% reduction in early marriage among young women. Similarly, our 2SLS estimates for childbearing using the Young Lives data were qualitatively large. Entering Grade 1 late reduced the probability of having a child by 25.3 percentage points among compliers of the school-entry age policy (i.e., those who were eligible to enter school late and complied with the policy), although the sample size is much smaller compared to the Census and confidence intervals are wide. These findings may point public health campaigns to prevent adolescent pregnancy and early marriage to young-for-grade students who may face higher risks. Flexible school entry policies may accommodate individual needs and support programs for younger students could be provided to maximize their educational engagement and health. Policymakers could also further integrate educational and sexual health interventions into the school curriculum, targeting students who are younger than their peers, to provide them with the knowledge and resources needed to make informed decisions about their future. For example, by providing information on the benefits of staying in school longer and the risks of adolescent pregnancy and marriage to young-for-grade students, which has been shown to be effective elsewhere ^36^.

This study has some limitations. First, respondents in the Young Lives Study were purposively sampled and, as with most detailed longitudinal studies, is limited in geographical scope ^23^. To mitigate potential concerns of selection bias and external generalizability, however, we sought to triangulate our results from the Young Lives data with data from three decades of Census data from Vietnam. Our results are generally consistent across different datasets and different outcome measures. Second, our results apply only to respondents up until ages 23 (but not beyond). We do not know whether the reductions observed in early family formation are “true” reductions in childbearing or whether adolescents merely postponed childbearing to a later date. A promising avenue for population health research would be exploit the natural experiment and data outlined in the current analysis to determine impacts longer term impacts of school-entry age ^37^ and implications of school-entry rules among parents for the health of the next generation ^38^. Third, our results apply only to the analyzed education policy in the context of Vietnam. The government of Vietnam exercises relatively tight control over development strategies and the responses of households and teachers to school-entry age policies may differ in other settings.

## Conclusion

This study reveals a compelling association between school-entry age and school attendance and family formation in adolescence in Vietnam. Children who enter school later tend to have more years of pre-primary education, stay in school longer, and postpone early family formation compared to their earlier-starting peers. Relative age for grade could be considered when designing sexual and reproductive health interventions and programs targeted to adolescents.

## Data Availability

All data produced in the present work are contained in the manuscript, and any additional data upon reasonable request will be made available.

https://beta.ukdataservice.ac.uk/datacatalogue/series/series?id=2000060#!/access-data

## Acknowledgements

We thank study participants and staff of the Young Lives research project and the Population and Housing Census in Vietnam. We also acknowledge the statistical offices that provided the underlying Census data which made this research possible, including the Bureau of the Central Steering Committee, General Statistics Office, Vietnam. We are also grateful to Caine Rolleston, Cuong Nguyen Viet, Jo Boyce, Maria Molina, Dani Shahzada, and Gary Rotto for helpful comments on this research project. The data come from Young Lives (www.younglives.org.uk) and IPUMS International (https://international.ipums.org/international/). The Young Lives project has been core-funded by UK aid from the Department for International Development (DFID). The views expressed here are those of the authors. They are not necessarily those of, or endorsed by, Young Lives, the University of Oxford, DFID or other funders.

## Appendix

### Text S1. Additional information on data sources

#### Young Lives Study

Data on socio-demographic characteristics, schooling, and health outcomes were extracted from Young Lives data. These datasets contain information on a longitudinal study of poverty and inequality that has been following the lives of 3,000 children in Vietnam over a 15-year period, surveyed once every 3-4 years since 2001 (Favara M et al, 2021). Round 1 of the study surveyed two groups of children, 1 year old (born in 2001-02) and 5 years old (born 1994-1995). Round 5 surveyed them when they were between 15 and 23 years old. The younger children were tracked from infancy to their mid-teens and the older children through into adulthood, when some became parents themselves. Data was collected from families, communities, schools, and directly from the children themselves. The Young Lives study is not intended to be a nationally representative survey but intends to show the impact of earlier circumstances (such as school-entry age) on children’s later educational and health outcomes. Multistage purposive sampling was used for sample selection, with the first stage involving a selection of 20 sentinel sites. The 20 sites are located in five provinces (Ben Tre, Da Nang, Hung. Yen, Lao Cai and Phu Yen). Households in selected sites were then chosen at random.

Attrition rates in the Young Lives data are low and over 90% of children were included in all rounds (rounds 1 to 5). Information on exact date of birth was classified as protected personal data and therefore not available. However, two other variables allow researchers to estimate month of birth. We thus estimated month of birth based on the date of interview (variable *dint*) and child’s age in months (variable *agemon*) which are both available in the dataset. The Young Lives Study estimated participant’s age in months by taking the age of the participant in days (date of interview minus date of birth) and dividing this number by 365/12 (number of days per month). Key advantages of the Young Lives data include the availability of data on school-entry age (i.e., age at beginning of Grade 1) and the ability to link school-entry age to measured educational and health outcomes up until early adulthood. Round 6 was a phone survey without anthropometric measurements and therefore not included in our analysis.

#### Vietnam Population and Housing Census

Data were extracted from the Vietnam Population and Housing Censuses of 1989, 1999, and 2009 through the Integrated Public Use Microdata Series (IPUMS). The Censuses were conducted by the Bureau of the Central Steering Committee, General Statistics Office, Vietnam, using a systematic stratified sampling to create random 5% (Census 1989) and 3% (Census 1999), and 15% (Census 2009) samples of the population universe. We limited the sample to all respondents who were born 3 months before and after the school-entry age cutoff (Dec 31^st^) to maximize the comparability of early starters and late starters. Data on month of birth, demographics, school attendance, childbearing, and marital status were available for 99% of eligible respondents ages 15-23 years, yielding a total sample of 1,595,365 individuals born between October and March. IPUMS harmonizes variables across Censuses so that the same codes have the same meaning across all Censuses.

The 1989 Census covered all residents in Vietnam, including those usually resident in Vietnam, but who were overseas at the time of the Census; special groups, including the police force, army and foreign affairs (de jure). Census day was April 1, 1989. The 1999 Census was conducted within 7 to 10 days of Census Day (April 1, 1999), and covered residents in Vietnam, including those usually resident in Vietnam, but who were overseas at the time of the census; special groups, including the police force, army and foreign affairs (de jure). The 2009 Census was conducted within 7 to 14 days of Census Day (April 1, 2009), and similarly covered residents in Vietnam, including those usually resident in Vietnam, but who were overseas at the time of the census; special groups, including the police force, army and foreign affairs (de jure).

The surveys provide information on demographic outcomes among all respondents, educational outcomes among respondents aged 5 years and older, and childbearing outcomes among women aged 15-49 years. The 1989, 1999, and 2009 Census contain data on month of birth based on the survey questions “Month and year of birth” [solar calendar month] (Census 1989), “In what solar calendar month and year was (Name) born?” (Census 1999), “In what solar calendar month and year was [the respondent] born?” (Census 2009) and was asked from all respondents. IPUMS harmonizes variables across surveys so that the same codes have the same meaning across all surveys. Data on month of birth was not available in the Vietnam Population and Housing Census of 2019 and therefore not used in the current analysis.

### Text S2. Education context in Vietnam

#### Education system

The Education Law of 1995 (Viet Nam National Assembly) describes the basic structure of the education system in Vietnam. The education system consists of five levels: preschool, primary school, secondary school, high school, and higher education. Basic education consists of five years of primary education, four years of secondary education, and three years of high school education. The government of Vietnam exercises tight control over development strategies. An increase in private sector engagement and transition to a market economy has brought in the need and focus to modernize education system by bringing in new reforms. Guided by the principle that an investment in education is an investment in economic development, the government has pursued reforming education by means of a highly competitive curriculum.

There is also a model of where reforms in the education system are implemented on a selective basis targeting large cities, economic priority zones, and urban areas. Various education reforms and strategies are developed and implemented over the period since the economic renovation commencing in 1986, educational reforms taken by the governments in 1994 to make education system more responsive to the labor market demands and requirements. In late 1997 the Minister of Education and Training announced a framework to modernize the education system to align with market reforms. The reforms aim to balance continuity with change, preserving cultural values while adapting to the demands of the modern global economy. Vietnam’s government has devoted between 15-20% of its entire spending budget to education since the later 1990s.

Enrollment in primary and secondary levels have been increasing since the early 1990s. Net primary rate of enrollment has increased from 85.6% in 1992-93 to 93.7% in 1997 and increased to 98.0% by 2014. In the span of 14 years, from 1992-93 to 2006 the children (in the age group of 15 -17) enrolled in the upper secondary school increased from one out of four to three out of four. Vietnam is committed to Education for All (EFA) and aligns with UNESCO’s goals. In 1990 Vietnam has endorsed the Jomtien Declaration, this declaration set the foundation for subsequent educational policies and reforms in Vietnam. At the World Education Forum in 2000 held in Dakar, Senegal, Vietnam has reaffirmed its commitment to EFA. The Dakar Framework emphasized six key goals, including universal primary education, gender equality, adult education, and quality of education. The report published by UNESCO in 2002 provides insight into Vietnam’s progress and challenges. Vietnam has also participated in the EFA Fast Track Initiative (Global Partnership of Education) launched by the World Bank in 2002 to help low-income countries accelerate progress towards EFA goal of enrolling all primary school-aged children in school by 2015. While Vietnam’s education system has made significant progress, challenges remain. While there is no gender gap at primary school level, from lower-secondary age group (ages 11 – 14) a gender gap begins to appear and increases in upper-secondary school level. There’s also a gap in urban and rural education. Ongoing reforms, investments, and efforts to address disparities are shaping the education landscape.

#### School-entry age policy

The Education Law of 1998 (Viet Nam National Assembly) also requires that a child enroll in Grade 1 of primary school at the start of the academic year in the calendar year they become 6 years old. Since eligibility is based on calendar year, month of birth *within* a year of birth does not matter for grade 1 eligibility as opposed to month of birth *across* years of birth. A child born December in year of birth X, for example, is eligible whereas a child born in January in year of birth X+1 may not be eligible. Specifically, article 22 of Section 1 of the Education Law of 1998 stipulates that: “Primary education is the compulsory level of education for all children from six to fourteen years old; it is conducted in five school years from the first to the fifth form. The age of pupils admitted to the first form is six years”. (Education Law of 1998) Similarly, the Education Law of 2019 also stipulates that “the entry age for the first grade is 6”. The school year starts in the first week of September and runs until the end of May the following calendar year. Primary schooling is compulsory and universal primary education has been achieved. As a result of the school-entry age policy, children who are born just before December 31^st^ start school one year earlier compared to children who are born just after December 31^st^. The policy has been suggested to be in place since 1945 when the nation became independent, although implementation and enforcement of the policy may have evolved over time.

**Figure S1.**
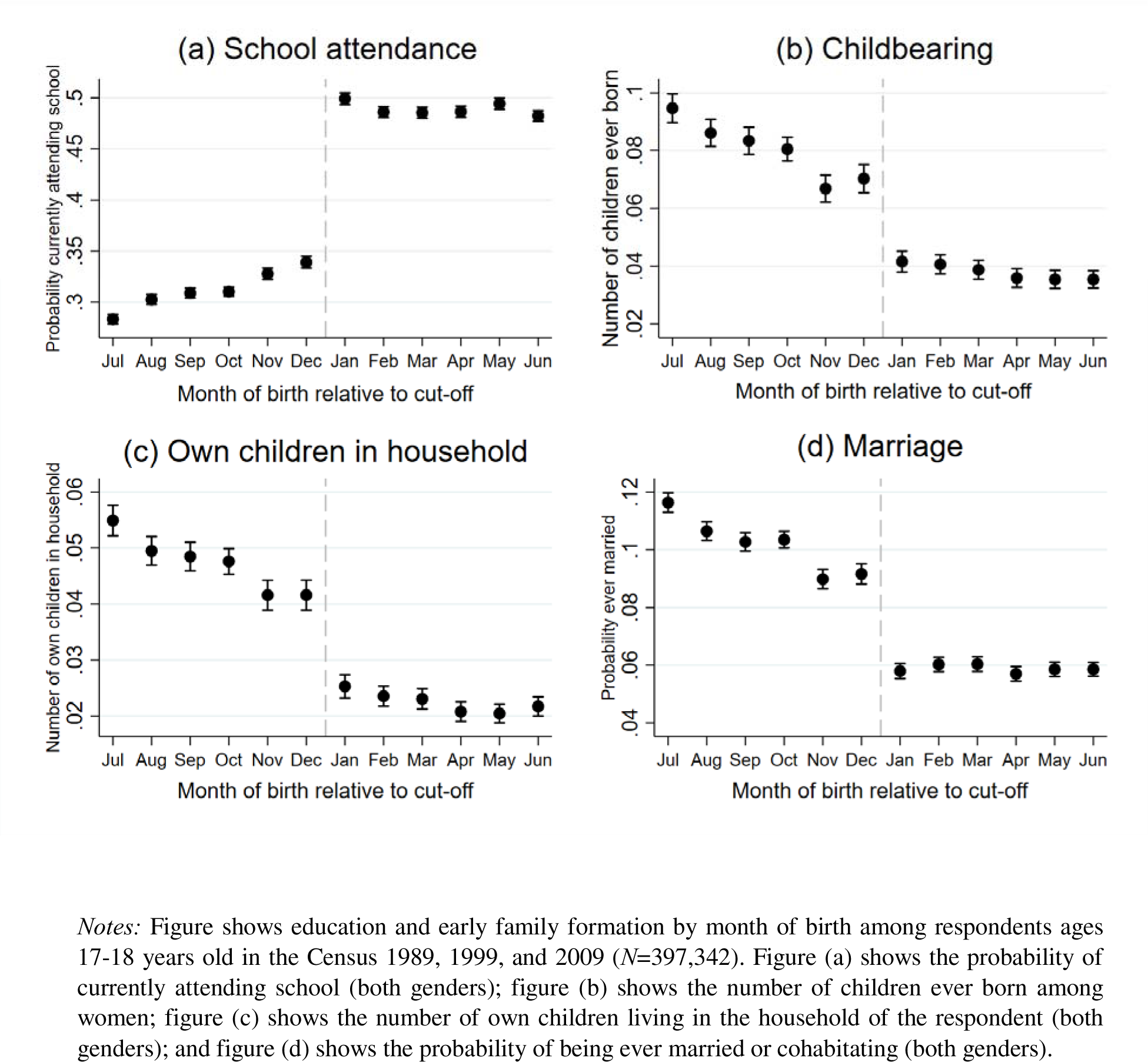
Adolescent childbearing and marriage by month of birth.

**Figure S2.**
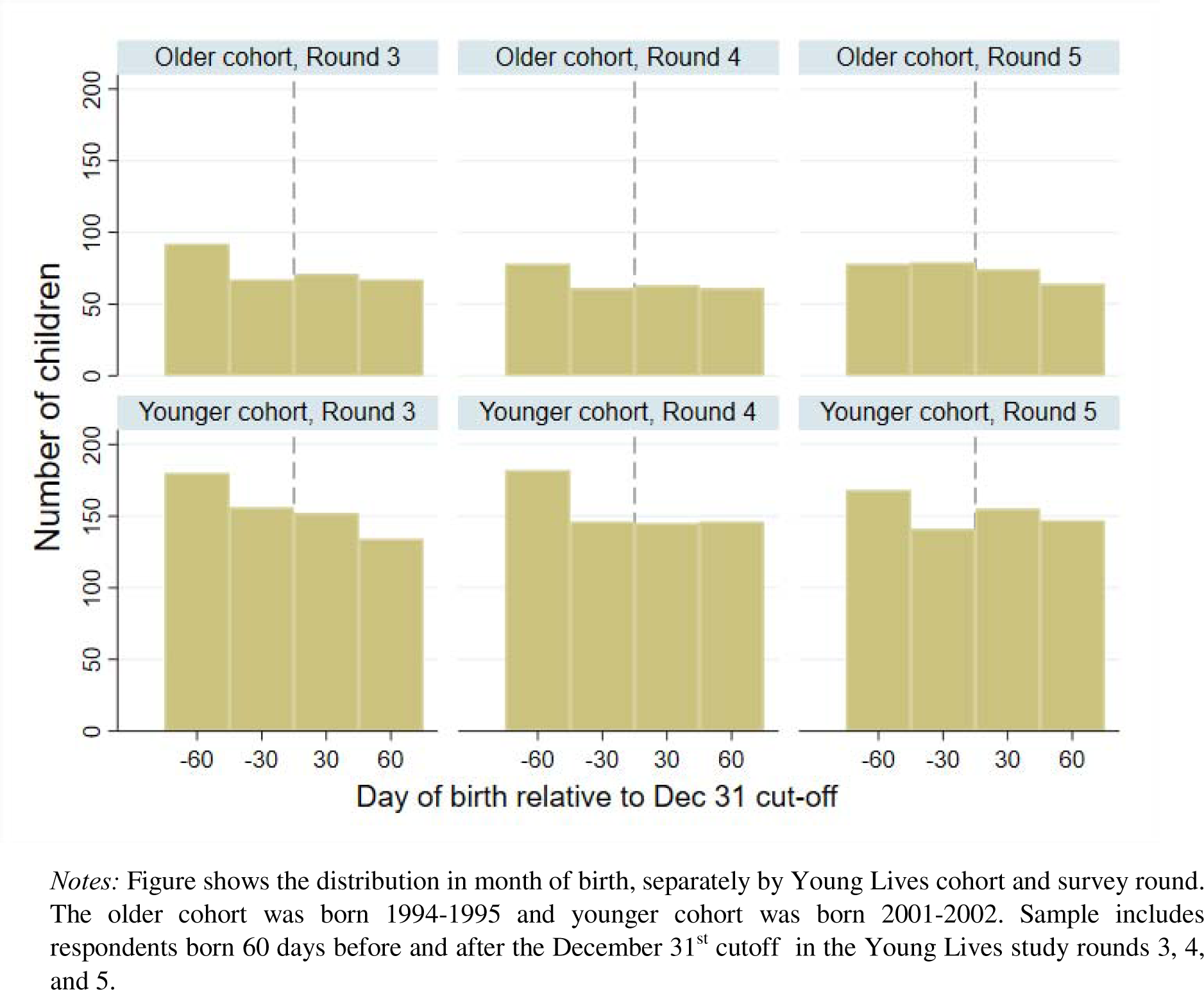
Distribution in month of birth by cohort and survey round.

**Figure S3.**
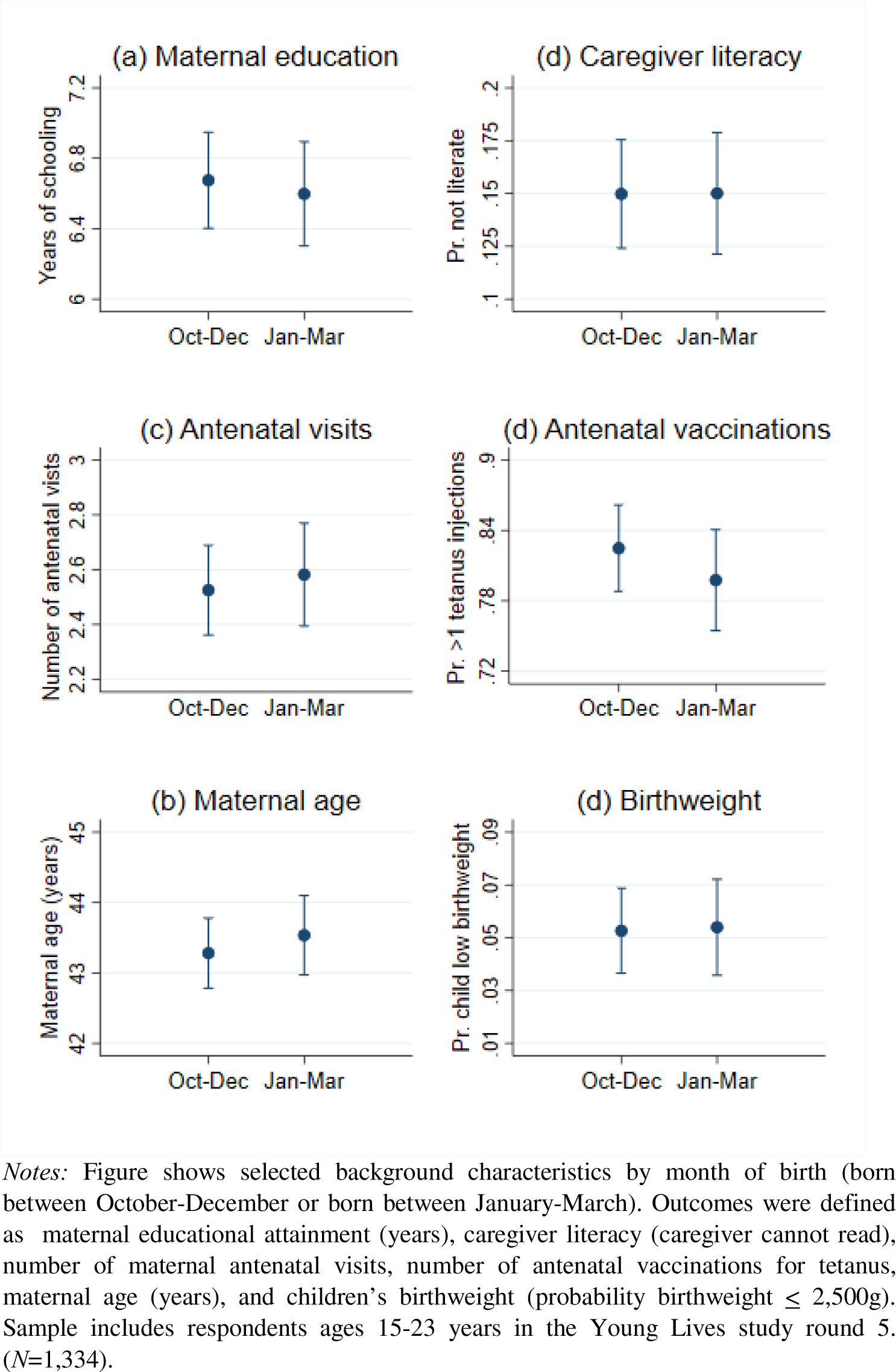
Balance in background characteristics.

**Figure S4.**
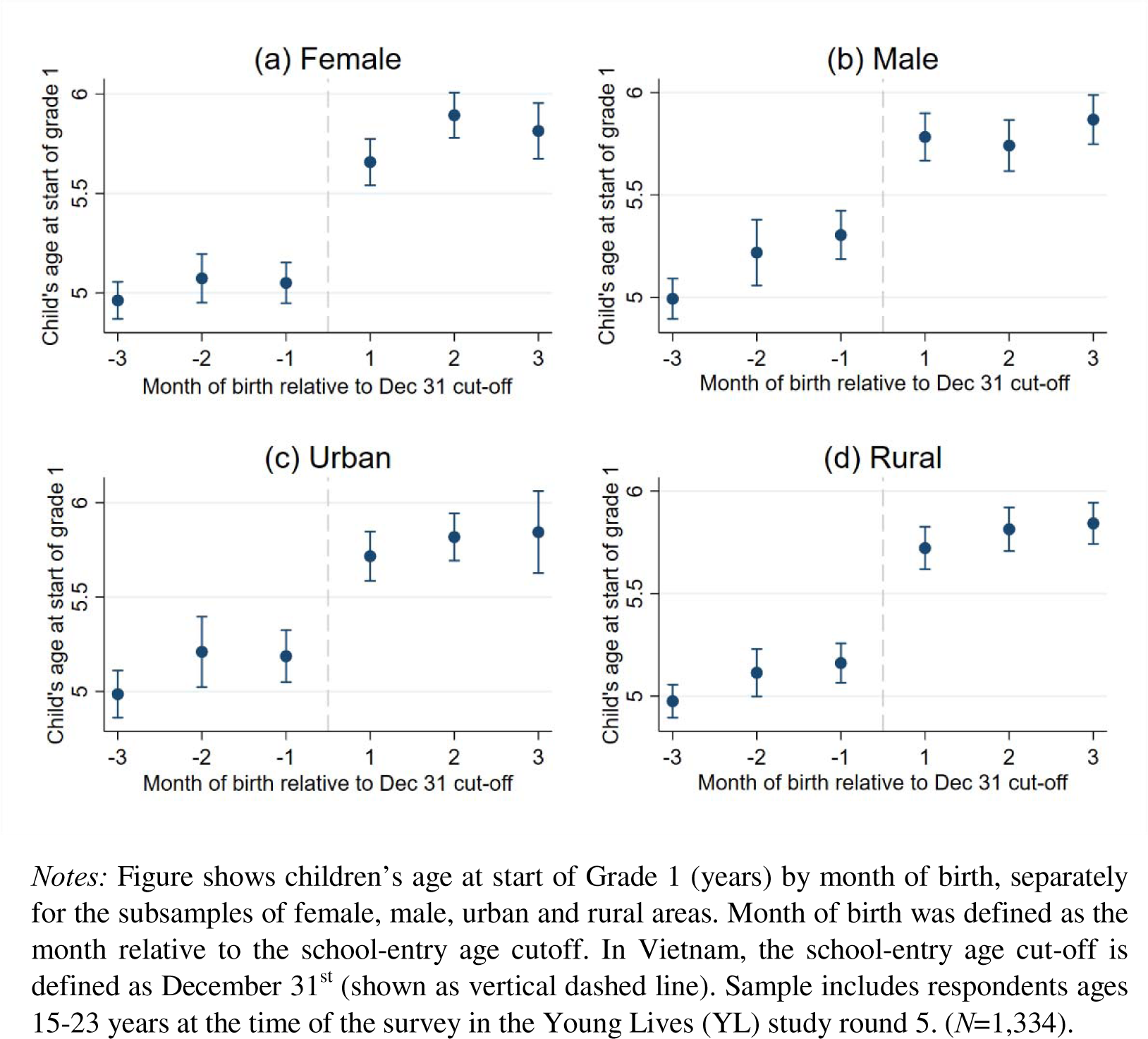
School-entry age and month of birth by gender and area.

**Table S1.** Selected characteristics of Young Lives sample.

| *Characteristic* | **Female** | **Male** | **Both genders** |
| --- | --- | --- | --- |
| *Age (years)* | 17.3 | 17.3 | 17.3 |
| *Area of residence, %* |  |  |  |
| Urban | 15.5 | 14.1 | 14.8 |
| Rural | 84.5 | 85.9 | 85.2 |
| *School-entry age (years)* | 5.3 | 5.4 | 5.4 |
| *Ever attended pre-school, %* |  |  |  |
| Yes | 68.3 | 71.4 | 69.9 |
| No | 31.7 | 28.6 | 30.1 |
| *Child is currently enrolled, %* |  |  |  |
| Yes | 65.8 | 61.7 | 63.8 |
| No | 34.2 | 38.3 | 36.2 |
| *Child's time spent in school (hours/day)* | 4.0 | 3.8 | 3.9 |
| *Child's health in general, %* |  |  |  |
| Very poor | 0.2 | 0.3 | 0.2 |
| Poor | 2.5 | 2.3 | 2.4 |
| Average | 68.8 | 58.1 | 63.5 |
| Good | 26.5 | 33.4 | 29.9 |
| Very good | 2.1 | 5.9 | 4.0 |
| *Any health issue since last round, %* |  |  |  |
| Yes | 17.8 | 19.5 | 18.6 |
| No | 82.3 | 80.1 | 81.2 |
| *Body mass index (kg/m<sup>2</sup>)* | 19.5 | 19.6 | 19.5 |
| Observations | 676 | 658 | 1,334 |

*Notes:* Sample includes respondents ages 15-23 years at the time of the survey in the Young Lives study Round 5 of Vietnam with complete data on educational outcomes and body-mass index and born between October and March. ( $N=1,334$ ).

**Table S2.**
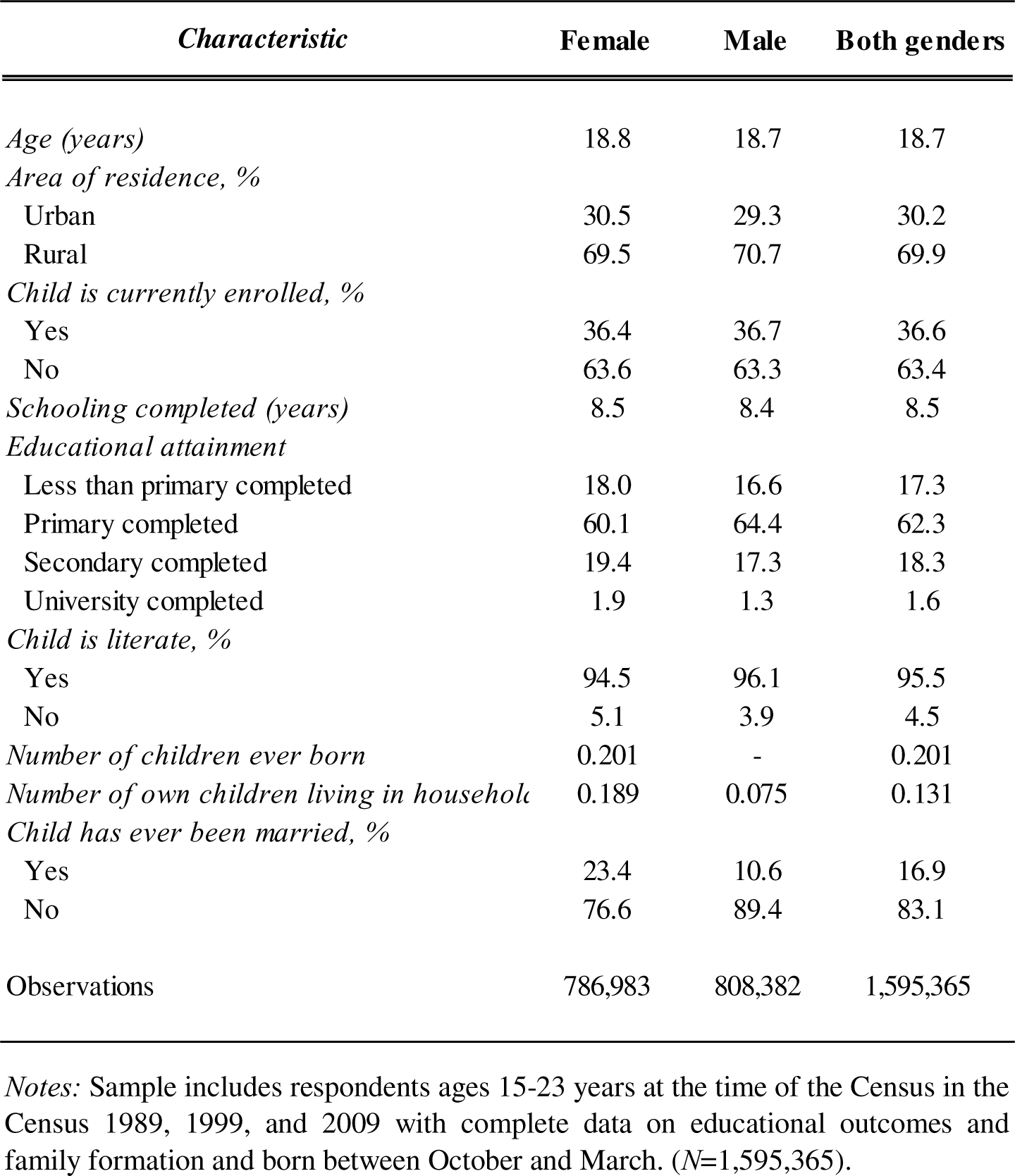
Selected characteristics of Census sample.

| *Characteristic* | **Female** | **Male** | **Both genders** |
| --- | --- | --- | --- |
| *Age (years)* | 18.8 | 18.7 | 18.7 |
| *Area of residence, %* |  |  |  |
| Urban | 30.5 | 29.3 | 30.2 |
| Rural | 69.5 | 70.7 | 69.9 |
| *Child is currently enrolled, %* |  |  |  |
| Yes | 36.4 | 36.7 | 36.6 |
| No | 63.6 | 63.3 | 63.4 |
| *Schooling completed (years)* | 8.5 | 8.4 | 8.5 |
| *Educational attainment* |  |  |  |
| Less than primary completed | 18.0 | 16.6 | 17.3 |
| Primary completed | 60.1 | 64.4 | 62.3 |
| Secondary completed | 19.4 | 17.3 | 18.3 |
| University completed | 1.9 | 1.3 | 1.6 |
| *Child is literate, %* |  |  |  |
| Yes | 94.5 | 96.1 | 95.5 |
| No | 5.1 | 3.9 | 4.5 |
| *Number of children ever born* | 0.201 | - | 0.201 |
| *Number of own children living in household* | 0.189 | 0.075 | 0.131 |
| *Child has ever been married, %* |  |  |  |
| Yes | 23.4 | 10.6 | 16.9 |
| No | 76.6 | 89.4 | 83.1 |
| Observations | 786,983 | 808,382 | 1,595,365 |

*Notes:* Sample includes respondents ages 15-23 years at the time of the Census in the Census 1989, 1999, and 2009 with complete data on educational outcomes and family formation and born between October and March. (N=1,595,365).

**Table S3.** ITT results using alternative windows around the eligibility cut-off.

|  | (1) | (2) | (3) | (4) | (5) | (6) |
| --- | --- | --- | --- | --- | --- | --- |
| *Dependent variable (DV):* | **Child has**<br>**attended pre-**<br>**primary school**<br>**(1=yes)** | **Currently**<br>**attending**<br>**school**<br>**(1=yes)** | **Hours per**<br>**spent day**<br>**in school** | **Has son or**<br>**daughter**<br>**(1=yes)** | **Ever**<br>**married or**<br>**cohabited**<br>**(1=yes)** | **Body mass**<br>**index**<br>**(kg/m2)** |
| *Intention-to-treat results* |  |  |  |  |  |  |
| *Panel A. Using smaller window of months of birth around cut-off (2 months)* |  |  |  |  |  |  |
| *Predictor* |  |  |  |  |  |  |
| Born January - February (1=yes, 0=no) | 0.093***<br>(0.023) | 0.119***<br>(0.025) | 0.732***<br>(0.171) | -0.088**<br>(0.044) | -0.084*<br>(0.051) | 0.137<br>(0.202) |
| *Additional covariates* |  |  |  |  |  |  |
| Age (years) | ✓ | ✓ | ✓ | ✓ | ✓ | ✓ |
| Gender | ✓ | ✓ | ✓ | ✓ | ✓ | ✓ |
| *Sample* |  |  |  |  |  |  |
| Born 2001-2002 | ✓ | ✓ | ✓ | - | - | ✓ |
| Born 1994-1995 | ✓ | ✓ | ✓ | ✓ | ✓ | ✓ |
| Observations | 906 | 906 | 906 | 295 | 295 | 906 |
| R-squared | 0.428 | 0.406 | 0.369 | 0.059 | 0.092 | 0.055 |
| *Panel B. Using larger window of months of birth around cut-off (5 months)* |  |  |  |  |  |  |
| *Predictor* |  |  |  |  |  |  |
| Born January - May (1=yes, 0=no) | 0.084***<br>(0.015) | 0.090***<br>(0.018) | 0.527***<br>(0.122) | -0.078**<br>(0.038) | -0.051<br>(0.042) | 0.190<br>(0.149) |
| *Additional covariates* |  |  |  |  |  |  |
| Age (years) | ✓ | ✓ | ✓ | ✓ | ✓ | ✓ |
| Gender | ✓ | ✓ | ✓ | ✓ | ✓ | ✓ |
| *Sample* |  |  |  |  |  |  |
| Born 2001-2002 | ✓ | ✓ | ✓ | - | - | ✓ |
| Born 1994-1995 | ✓ | ✓ | ✓ | ✓ | ✓ | ✓ |
| Observations | 1,958 | 1,958 | 1,958 | 630 | 630 | 1,958 |
| R-squared | 0.498 | 0.387 | 0.356 | 0.054 | 0.07 | 0.038 |

*Notes:* Table shows Intention-to-treat (ITT) ordinary least squares (OLS) linear probability models. All models controlled for age (continuously in single years) and gender. Sample includes respondents ages 15-23 years at the time of the survey in the Young Lives study Round 5 of Vietnam. Data on fertility and marriage was not available for the younger birth cohorts in the Young Lives study. \*\*\* p<0.01, \*\* p<0.05, \* p<0.1

**Table S4.** ITT results separately by Young Lives Survey round (2009-2016)

|  | (1) | (2) | (3) | (4) |
| --- | --- | --- | --- | --- |
| *Dependent variable (DV):* | **Child has**<br>**attended pre-**<br>**primary school**<br>**(1=yes)** | **Currently**<br>**attending**<br>**school**<br>**(1=yes)** | **Hours per**<br>**spent day**<br>**in school** | **Body mass**<br>**index**<br>**(kg/m2)** |
| *Intention-to-treat results* |  |  |  |  |
| *Panel A. Using Young Lives Round 3 (2009-2010)* |  |  |  |  |
| *Predictor* |  |  |  |  |
| Born January - March (1=yes, 0=no) | 0.065***<br>(0.019) | 0.028**<br>(0.014) | 0.244***<br>(0.094) | -0.067<br>(0.161) |
| *Additional covariates* |  |  |  |  |
| Age (years) | ✓ | ✓ | ✓ | ✓ |
| Gender | ✓ | ✓ | ✓ | ✓ |
| Mean DV, October - DececeMBER birth cohorts | 0.685 | 0.911 | 4.7 | 16.1 |
| Observations | 1,351 | 1,351 | 1,351 | 1,351 |
| R-squared | 0.441 | 0.151 | 0.049 | 0.179 |
| *Panel B. Using Young Lives Round 4 (2013)* |  |  |  |  |
| *Predictor* |  |  |  |  |
| Born January - March (1=yes, 0=no) | 0.051***<br>(0.019) | 0.035**<br>(0.017) | 0.152<br>(0.117) | -0.014<br>(0.151) |
| *Additional covariates* |  |  |  |  |
| Age (years) | ✓ | ✓ | ✓ | ✓ |
| Gender | ✓ | ✓ | ✓ | ✓ |
| Mean DV, October - DececeMBER birth cohorts | 0.717 | 0.818 | 4.7 | 17.8 |
| Observations | 1,309 | 1,309 | 1,309 | 1,309 |
| R-squared | 0.407 | 0.411 | 0.269 | 0.201 |
| *Panel C. Using Young Lives Round 5 (2016)* |  |  |  |  |
| *Predictor* |  |  |  |  |
| Born January - March (1=yes, 0=no) | 0.090***<br>(0.019) | 0.113***<br>(0.020) | 0.632***<br>(0.144) | 0.075<br>(0.163) |
| *Additional covariates* |  |  |  |  |
| Age (years) | ✓ | ✓ | ✓ | ✓ |
| Gender | ✓ | ✓ | ✓ | ✓ |
| Mean DV, October - DececeMBER birth cohorts | 0.669 | 0.599 | 3.7 | 19.5 |
| Observations | 1,334 | 1,334 | 1,334 | 1,334 |
| R-squared | 0.452 | 0.392 | 0.359 | 0.062 |

*Notes:* Table shows Intention-to-treat (ITT) ordinary least squares (OLS) linear probability models. All models controlled for age (continuously in single years) and gender. Sample includes respondents in the Young Lives study Round 3 (ages 7 – 16 years), Round 4 (ages 12 – 20 years), and Round 5 (ages 15 – 23 years). Data on fertility and marriage was only available for later rounds among the older birth cohorts in the Young Lives study. \*\*\* p<0.01, \*\* p<0.05, \* p<0.1

